# Exploring Epidemiological Behavior of Novel Coronavirus Outbreak through the Development and Analysis of COVID-19 Daily Dataset in Bangladesh

**DOI:** 10.1101/2020.06.30.20143909

**Authors:** Samrat Kumar Dey, Md. Mahbubur Rahman, Umme Raihan Siddiqi, Arpita Howlader

**Affiliations:** Department of Computer Science and Engineering (CSE), Dhaka International University (DIU), Dhaka-1205, Bangladesh; Department of Computer Science and Engineering (CSE), Military Institute of Science and Technology (MIST), Mirpur Cantonment, Dhaka-1216, Bangladesh; Department of Physiology, Shaheed Suhrawardy Medical College (ShSMC), Dhaka-1207, Bangladesh; Department of Computer and Communication Engineering (CCE), Patuakhali Science and Technology University (PSTU), Dumki-8602, Patuakhali, Bangladesh

**Keywords:** COVID-19 BD Dataset, outbreak, data analysis, coronavirus, visualization, SARS-CoV-2, Bangladesh

## Abstract

**Background:** Globally, there is an obvious concern about the fact that the evolving 2019-nCoV coronavirus is a worldwide public health threat. The appearance of severe acute respiratory syndrome coronavirus 2 (SARS-CoV-2) in China at the end of 2019 triggered a major global epidemic, which is now a major community health issue. As of April 17, 2020, according to Institute of Epidemiology, Disease Control and Research (IEDCR) Bangladesh has reported 1838 confirmed cases in between 8 March to 17 April 2020, with > 4.08% of mortality rate and >3.15% of recovery rate. COVID-19 outbreak is evolving so rapidly in Bangladesh; therefore, the availability of epidemiological data and its sensible analysis are essential to direct strategies for situational awareness and intervention.

**Method:** This article presents an exploratory data analysis approach to collect and analyze COVID-19 data on epidemiological outbreaks based on first publicly available COVID-19 Daily Dataset of Bangladesh. Various publicly open data sources on the outbreak of COVID-19 provided by the IEDCR, World Health Organization (WHO), Directorate General of Health Services (DGHS), and Ministry of Health and Family Welfare (MHFW) of Bangladesh have been used in this research.

**Results:** A Visual Exploratory Data Analysis (V-EDA) techniques have been followed in this research to understand the epidemiological characteristics of COVID-19 outbreak in different districts of Bangladesh in between 8 March 2020 to 12 April 2020 and these findings were compared with those of other countries.

**Conclusions:** In all, this is extremely important to promptly spread information to understand the risks of this pandemic and begin containment activities in the country.

## 1. Introduction

Bangladesh is a democratic nation surrounded by east, west and north India, Myanmar from southeast with Bay of Bengal on the southern side. As of 1 July 2017, the population size in Bangladesh is estimated at 162.7 million (male female ratio is 100.2:100). It has a unitary form of government, without a province or a territory. There are 64 districts in the country and again each district is divided into several sub districts (which is called upazilas). Right now, there are eight divisions (Barisal, Chittagong, Dhaka, Khulna, Mymensingh, Rajshahi, Rangpur, and Sylhet) in the country and each division is named after the major city within its jurisdiction, which also acts as the administrative headquarter of that division. The Ministry of Health and Family Welfare (MoHFW) is responsible for the preparation and management of curative, preventive and pro-active health services for the population of the country and therefore considered as one of the largest ministries of the Government of Bangladesh.

Institute of Epidemiology, Disease Control and Research (IEDCR) is considered as the focal institute for conducting public health surveillance and outbreak response. During the last few years, however, the efficiency of the International Health Regulation (IHR) has dramatically increased and exceeded 68% in the country.^**1**^ Moreover, it also has the national ability to test COVID-19 patients by using polymerase chain reaction (PCR) machine confirmed laboratory. The first cases of pneumonia of unknown origin identified in Wuhan City, the capital of Hubei Province, China in early December 2019 was later determined as a non-SARS novel coronavirus by the Chinese Center for Disease Control and Prevention (CDC).^**2**^ Coronaviruses are non-segmented positive-sense RNA viruses that belong to the *Coronaviridae* family and the *Nidovirales* order and are widely distributed in humans and other mammals.^**3**^ The pathogen has been described as a novel enveloped RNA *betacoronavirus*^**4**^ commonly referred to as severe acute respiratory coronavirus 2 syndrome (SARS-CoV-2), having a phylogenetic resemblance to SARS-CoV.^**5**^ With a mortality rate of 10% for SARS-CoV and 37% for MERS-CoV,^**6**,**7**^ the epidemics of the two beta coronaviruses, severe acute respiratory syndrome coronavirus (SARS-CoV), ^**8**,**9**^ and Middle East respiratory syndrome coronavirus (MERS-CoV),^**10**,**11**^ have caused more than ten thousand cumulative cases in the last two decades. With no surprise, on 30 January 2020, World Health Organization (WHO) declared the COVID-19 outbreak as a Public Health Emergency of International concern (PHEIC).^**12**^ As of April 15, 2020, a total of 1914916 laboratory-confirmed cases have been documented globally and among them, Bangladesh has confirmed 1132 cases with a death rate of 4.4% ^**13**^ .This report will investigate Bangladesh’s epidemiological data based on the country’s first COVID-19 daily dataset between March 8, 2020 and April 12, 2020.

Until 12 April 2020, Bangladesh has confirmed 621 laboratory tested positive patients affected by SARS-CoV-2 virus with 621 confirmed, 39 recovered, and 34 death, 548 infected, 9,653 tested cases. On February 5, 2020, IEDCR performed coronavirus tests on 39 Bangladeshis for the very first time; those came back Bangladesh from China on that time. Until then, none of them found infected with the virus. In these circumstances, turning public fears into reality, on 8 March 2020, Bangladesh has confirmed its first case of COVID-19 patients (N=3) caused by SARS-CoV-2 virus. Among these, two of the three infected were all aged between 20 and 35 and returned recently to Bangladesh from different cities in Italy. However, this is significant that, only after 7 days of first confirmed COVID-19 reported case, on 16 March 2020, IEDCR reported that the virus had begun to spread locally, and indicated that children in the country had been infected with coronavirus for the very first time. As the outbreak of COVID-19 is expanding rapidly globally, already appeared as a global pandemic, therefore we aim to describe the number of different cases caused by SARS-CoV-2 and represents the epidemiological data through visual exploratory data analysis (EDA)^**14**^ approach. Considering COVID-19’s rapid spread, we have designed a comprehensive daily dataset^**15**^ and decided that an updated case review with outbreak analysis in Bangladesh may help identify the epidemiological characteristics and severity of the disease across the country. Apart from preparing an inclusive dataset, this research will also analyze the different trend of documented cases of COVID-19 in the countries of the South-East Asia Region (SEAR), and South Asian Association for Regional Cooperation (SAARC). Moreover, this work would pose a concern for the citizens of Bangladesh, whether the situation of COVID-19 in the country is on the verge of a rapid outbreak in near future or if it can flatten the curve. As of April 16, 2020, according to the situation report provided by the IEDCR^**16**^, ten new death cases taking the total number of death cases to 60 in the country and conformed case raised to 1572.

## 2. Methods and Material

In this research, we have used numerous range of data sources for preparing our datasets. This dataset is named as “Bangladesh COVID-19 Daily Dataset” available at this URL: https://www.kaggle.com/samratkumardey/bangladesh-covid19-daily-dataset. As per our knowledge “Bangladesh COVID-19 Daily Dataset” is the first designed and developed dataset that enlist the different cases (confirmed, death, recovered, number of test etc.) of COVID-19 outbreak in Bangladesh. We use official government sources include (a) daily press briefing provided by IEDCR, (b) press releases from official website of Corona BD (https://corona.gov.bd/), (c) press releases on the official websites of Ministries of Health (http://www.mohfw.gov.bd/), and Directorate General of Health Services (DGHS) (https://dghs.gov.bd/index.php/en/). In order to prepare a comprehensive and precise dataset, we collected data on the following: (i) Daily dates, which include the date of daily new confirmed case, total confirmed case, daily new deaths, total deaths, daily new recovered, total recovered, daily new tests, total tests, and active cases. (ii) Geographic information, division and district wise confirmed cases. (iii) Demographic statistics about the patient’s age and sex. (iv) Date wise statistics of quarantine and isolated patients. Table 1 provides insight on Bangladesh COVID-19 Daily Dataset and their respective data files with their column description. As this is a time series data, therefore the number of instances on a given day is the cumulative number. This dataset contains different COVID-19 cases data from 8 March 2020 to 12 April 2020. It is to be notated that Bangladesh has familiarized with its very first laboratory tested confirmed COVID-19 patients on 8 March 2020 and it reported its first death case on 18 march, 2020. Our research team believes that these data should be available to policy makers, health professionals and data analysts and should be readily accessible.

**Table 1:** Tabular representation of Bangladesh COVID-19 Daily Dataset with columns description

| Bangladesh COVID-19 Daily Dataset |  |  |
| --- | --- | --- |
| Data files | Columns | Description |
| COVID-19 BD Dataset-10 april.csv | Date | Specific date |
|  | Daily new confirmed cases | Day wise new confirmed case reported |
|  | Total confirmed cases | Aggregate confirmed cases of each day |
|  | Daily new deaths | Day wise new death case reported |
|  | Total deaths | Aggregate death cases of each day |
|  | Daily new recovered | Day wise new recovered case reported |
|  | Total recovered | Aggregate recovered cases of each day |
|  | Daily New Tests | Day wise number of new test performed |
|  | Total Tests | Cumulative number of test of each day |
|  | Active Cases | Till date number of active patient |
| COVID-19 BD Isolation statistics.csv | Date | Specific date |
|  | Total Isolation | Cumulative isolated patients till date |
|  | Currently in Isolation | Number of isolation patients till now |
|  | Release from Isolation | Number of patients released from isolation |
| COVID-19 BD Quarantine statistics.csv | Date | Specific date |
|  | Total Quarantine | Cumulative quarantine persons till date |
|  | Currently in Quarantine | Number of quarantine persons till now |
|  | Release from Quarantine | Number of person released from quarantine |
| COVID 19 BD District wise confirmed cases.csv | Division | Name of the division |
|  | District | Name of the district |
|  | Number of confirmed cases till date | Cumulative case number |
| COVID 19 BD Dhaka | City /Location | Dhaka city location |
| location wise confirmed cases.csv | Number of confirmed cases till date | Aggregate confirmed cases of each day till date at Dhaka city |

## 3. Results

This exploratory data analysis (EDA) mainly focused on the COVID-19 outbreak of a South East Asian country, Bangladesh. Apart from designing and developing a dataset, this research also analyzed the different characteristics of COVID-19 epidemic in between 8 March 2020 to 12 April 2020. All the data analysis and comparative results are based on the data that we collected and curated individual-level data from national level, division level, district level, and official health reports provided by government officials, as well as additional information from WHO situation report. As of 12 April 2020, according to the Institute of Epidemiology, Disease Control and Research (IEDCR), there are 621 confirmed COVID-19 cases by rt-PCR in Bangladesh, including 39 patients who have recovered and 34 related deaths; Case Fatality Rate (CFR) is 05.48%. Figure 1 shows the daily distribution of confirmed, deaths, recovered and active COVID-19 cases from 08 March 2020 to 12 April 2020 in Bangladesh. Between 1 March 2020 and 12 April 2020, among the 621 COVID-19 reported cases, 69.9% (434) were males whereas 30.1% (187) were females. As of 12 April 2020, among the 621 COVID-19 reported cases, 88.2% (548/621) are currently under treatment, 6.3% (39) were cured and 5.5% (34) died. Distribution of confirmed COVID-19 cases by age, gender and division is shown on Table 2. Sixty percent (60%) (380/621) of all COVID-19 confirmed cases were between 21 and 50 years old, followed by 28% (63) of the age group 51 years and above, while the age group of less than 20 years represented only 10% (178) of all reported cases.

**Table 2:** Distribution of confirmed COVID-19 cases by Age, Gender and Division, 08 March – 12 April 2020 in Bangladesh

| Age distribution of Confirmed Patients |  |  |
| --- | --- | --- |
| Age group | Number of Cases | Percentage% |
| > 60 years | 17 | 2.737% |
| 51-60 years | 46 | 7.407% |
| 41-50 years | 122 | 19.645% |
| 31-40 years | 143 | 23.027% |
| 21-30 years | 115 | 18.518% |
| 11-20 years | 95 | 15.297% |
| < 10 years | 83 | 13.365% |
| Total | (N=621) | 100% |
| Gender distribution of Confirmed Patients |  |  |
| Sex | Number of Cases | Percentage % |
| Male | 434 | 69.9% |
| Female | 187 | 30.1% |
| Distribution of confirmed COVID-19 cases by Division |  |  |
| Division | Number of Cases | Percentage % |
| Barisal | 7 | 1.127% |
| Chittagong | 35 | 5.636% |
| Dhaka | 529 | 85.185% |
| Khulna | 1 | 0.161% |
| Mymensingh | 14 | 2.254% |
| Rajshahi | 0 | 0 |
| Rangpur | 15 | 2.415% |
| Sylhet | 3 | 0.483% |
| Unknown | 17 | 2.737% |
| Total | (N=621) | 100% |

**Figure 1:**
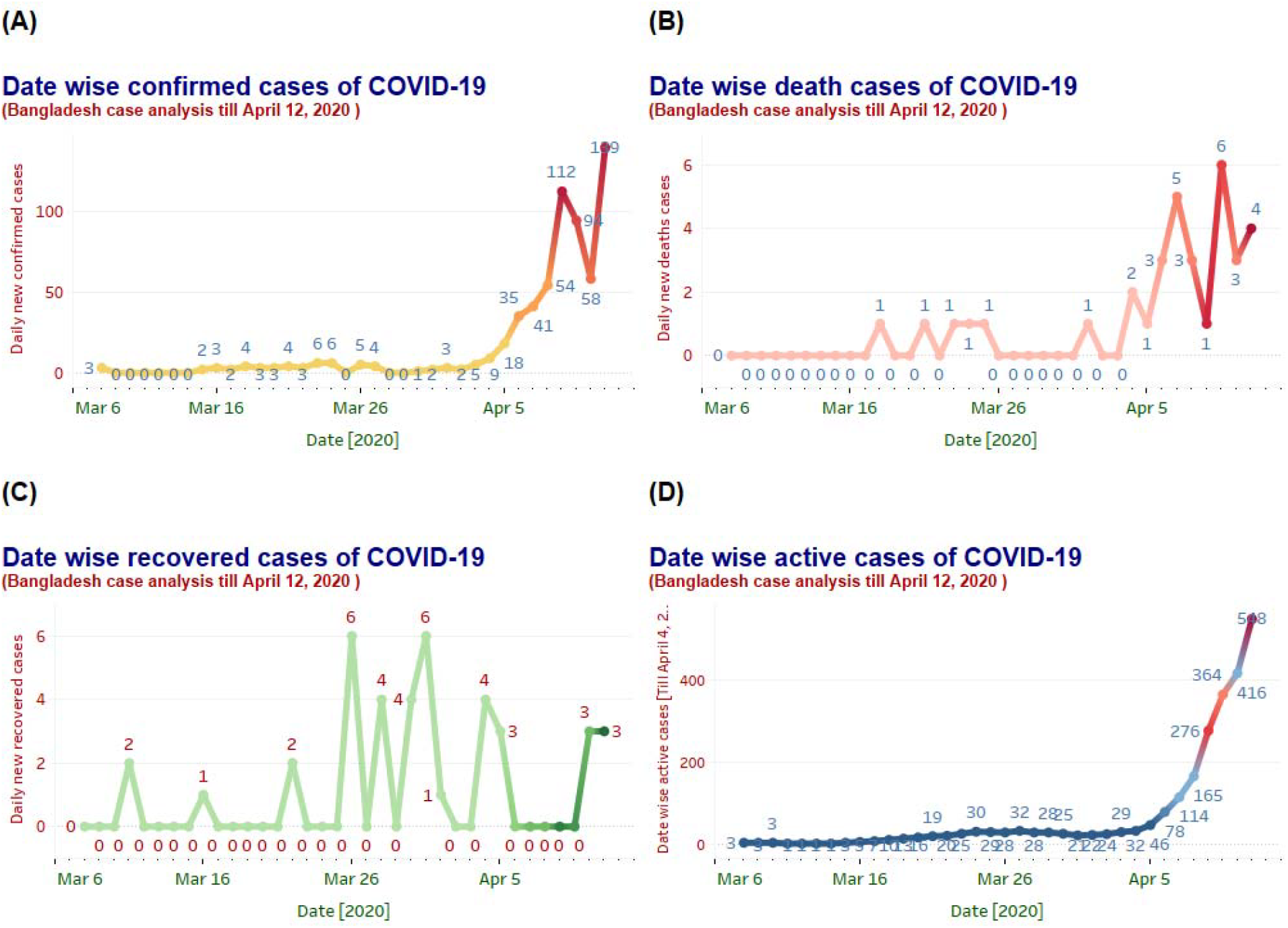
Graphs provide an overview of how COVID-19 is spreading in Bangladesh for the last 35 days (from 8 March 2020 to 12 April 2020). Until date, Bangladesh has confirmed 621 laboratories tested COVID-19 positive patients. 1(A) shows the specific date wise number of confirmed cases for each day. 1(B) presents the number of death case reported each day. Until 12 April 2020, Bangladesh has reported 34 (5.47%) deaths from 18 March 2020 (first death case reported). 1(C) and 1(D) enlist the date wis number of recovered patients 39 (6.28%) from COVID-19 and available patients under treatment 88.2% (548) until 12 April 2020.

It is evident that there has been an increasing trend in the number of reported confirmed cases since 3 April 2020, due to expansion of COVID-19 testing in the country and establishing testing capacity in 16 laboratories (Figure 2). By 12 April, a total of 9653 samples were tested of them 26.08% (2518) were tested by laboratories outside Dhaka city and 73.91% (7135) were tested by laboratories inside Dhaka. The overall COVID-19 test rate in Bangladesh is 8.3/100,000 population (until 12 April 2020). The Figure 1 shows the daily distribution of confirmed COVID-19 cases by Division, 08 March – 12 April 2020 in Bangladesh.

**Figure 2:**
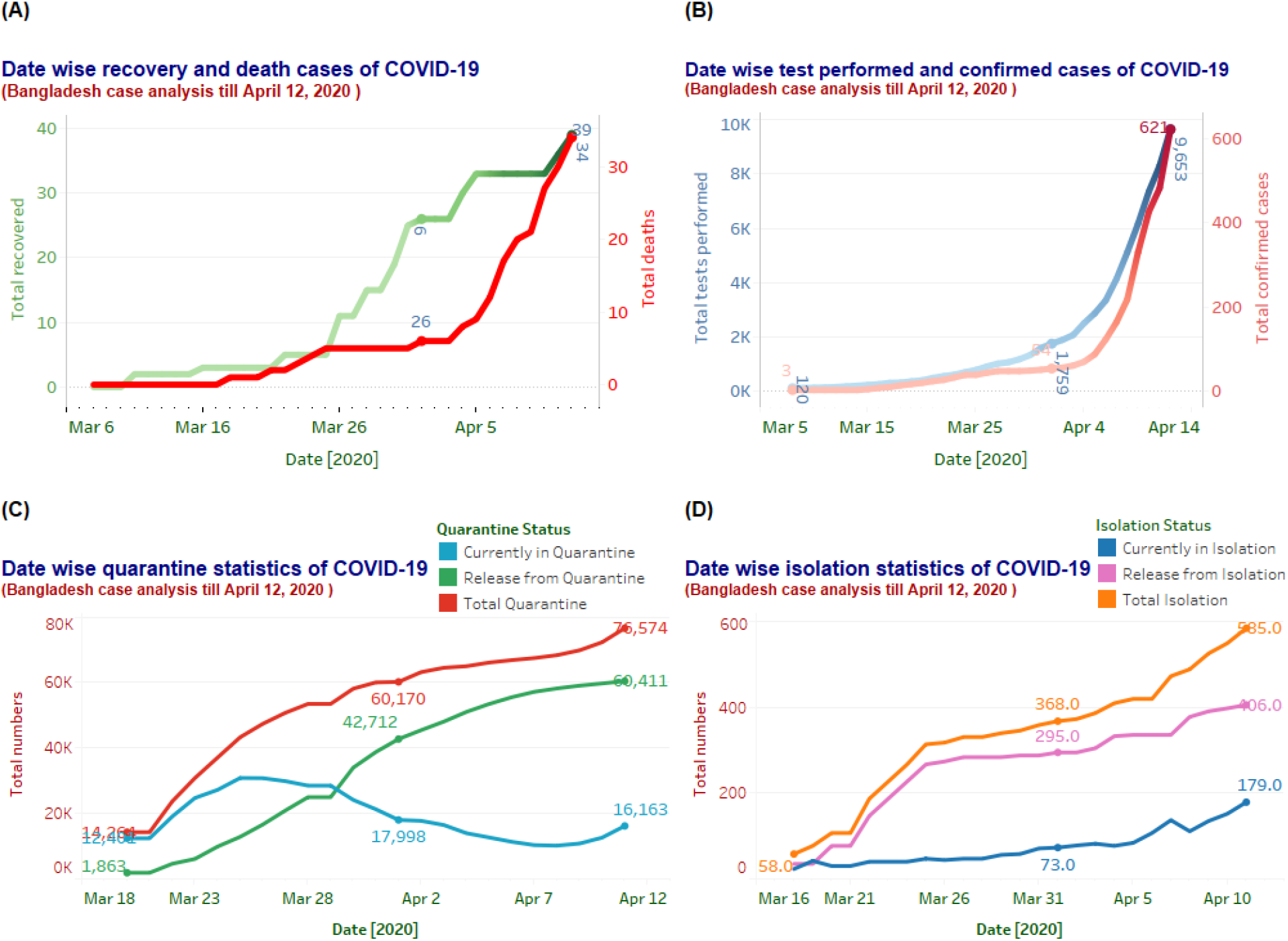
This graphical analysis will lead us to understand the current situation of COVID-19 outbreak in Bangladesh. 2(A) offers a comparative date wise representation of death cases with recoveries. Until 12 April 2020, Bangladesh has reported 5.47% (34) death cases, whereas 6.28% (39), patients recovered successfully among 621 confirmed patients. Another date wise comparison of the number of COVID-19 tests performed and the number of confirmed cases are shown on 2(B). With time, the number of the test-performed curve rises along with the total reported confirmed case. Around 9653 tests performed until 12 April 2020 and among them 621 patients found COVID-19 positive. 2(C) and 2(D) shows the date wise quarantine and isolation statistics for the last 35 days managed by the Government of Bangladesh.

Dhaka remains the area of the highest concentration of the reported COVID-19 cases (Table 2 & Figure 3). As of 12 April, 85% (529/621) of all confirmed cases were reported form Dhaka Division, followed by Chittagong division 6% (35/621), Rangpur division 3% (15/621), Mymensingh division 2.5% (14/621), Sylhet Division 0.5% (3/621)Barisal division 1% (7/621) and the remaining 0.5% (1/621) were reported from Khulna division. However, almost 3% confirmed cases are still unidentified in terms of their location.

**Figure 3:**
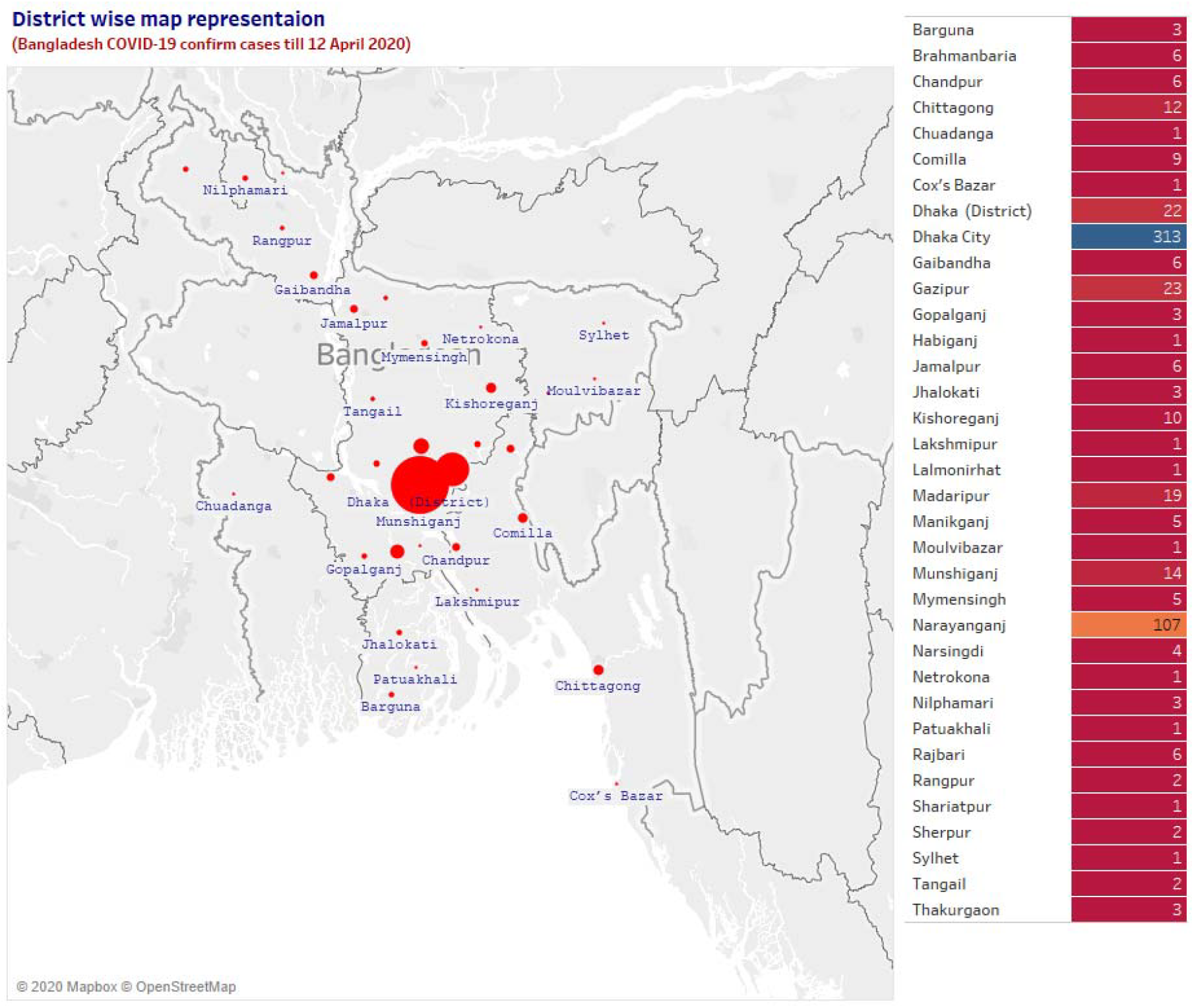
Distribution of COVID-19 laboratory tested confirmed cases in different district of Bangladesh from 8 March 2020 to 12 April 2020. Dhaka, the capital of the country shows the highest number of reported COVID-19 confirmed cases.

As of 12 April, there were 16163 individuals at quarantine (both in home and institutional quarantine); since home quarantine was established in the country to 12 April 2020, a total of 76574 individuals were quarantined of them 78.90% (60, 411) were released (Figure 2). Similarly, 179 individuals were then in isolation whereas a total of 59.03% (585) individuals were in isolation; among them 40.97% (406) were released from isolation till 12 April 2020. A date wise recovery with death cases analysis and number of test performed each day with the number of confirmed cases is also presented in this research (Figure 2).

Between 17 March 2020 to 12 April 2020, 76261 individuals were placed under home quarantine all the over the county, out of them 79% (60355) have been already released. Regarding isolation facility, as of 12 April, there are 7693 COVID-19 isolation beds in Bangladesh, of them 29% (22247/7693) are in Dhaka division, 16% (1200) in Rajshahi, 13% (1030) in Mymensingh, 12 % (848) in Chittagong, 10% (787) in Rajshahi, 8% (690) in Khulna, 7% (545) in Barisal, and 4% (346) in Sylhet.

In Dhaka division, Dhaka city reported 59% of the total confirmed cases for the division (313/529) followed by Narayanganj 20% (107), Gazipur 4% (23), Dhaka district 4% (22), Madaripur 4% (19), Munshigonj 3% (14) Kishoreganj 2% (10), the remining 4% (21) were reported from Rajbari (6), Manikganj (5) Narshingdi (4), Gopalganj (3), Tangail (2) and Shariatpur (1). Moreover, in Chittagong division, Chittagong City reported 34% of the total for the division (12/35), followed by Cumilla 26% (9), B. Baria 16% (6), Cox’s bazar 3% (1) and Laksmipur 3% (1).

Due to geographic orientation, Bangladesh enlisted as a country of South East Asian Region (SEAR) and it also work together with the South Asian Association for Regional Cooperation (SAARC) forum. Therefore, this research also compares the confirmed and mortality ratio in terms of days with these neighboring countries of the same region. However, this study also covers countries that had higher mortality rates worldwide until April 12, 2020 (Figure 4). This study correlates Bangladesh’s with SEAR, SARRC and top 10 countries (based on the highest number of recorded deaths) in terms of the number of days it takes to show a similar trend based on confirmed and mortality case pattern.

**Figure 4:**
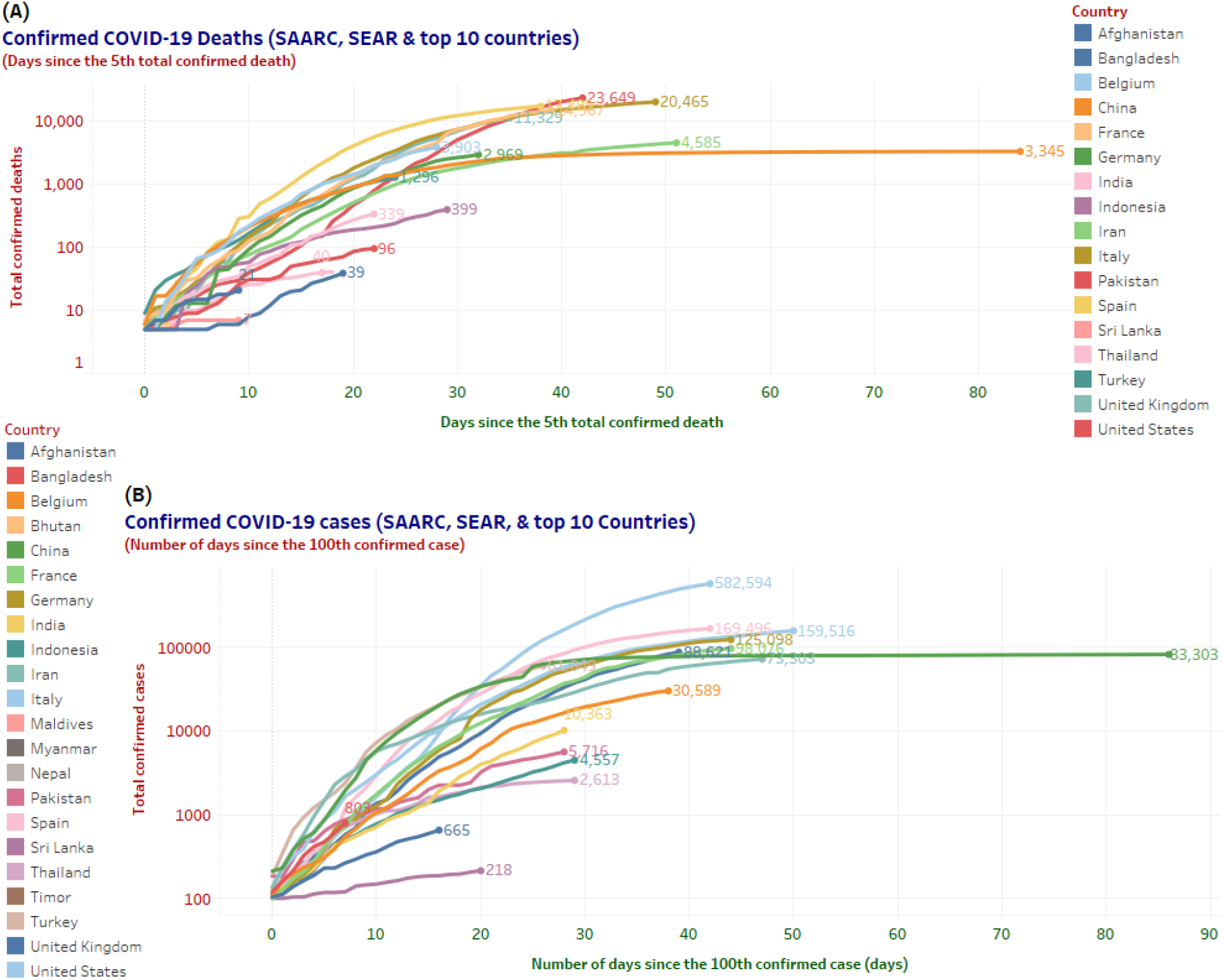
Comparative analysis in terms of days since the fifth total confirmed death and number of days since the 100^th^ confirmed cases with SAARC, SEAR and top 10 countries (highest death reported).

## 4. Discussion

We report here all the different cases of COVID-19 outbreak in Bangladesh based on daily dataset and compare its results with the countries of South Asian Association for Regional Cooperation (SAARC), South East Asian Region (SEAR) and globally top ten countries in terms of their daily death rate in between 8 March 2020 to 12 April 2020. Moreover, design and development of country’s first “Bangladesh COVID-19 Daily Dataset” is also focused in this research. Like other South Asian Countries, Bangladesh also outlined measures and plans to face this severe pandemic of COVID-19 with utmost importance. As of 15 April 2020, according to the Institute of Epidemiology, Disease Control and Research (IEDCR), there are 1231 confirmed COVID-19 cases in Bangladesh, including 49 patients who have recovered and 50 related deaths; Case Fatality Rate (CFR) is 04.06%. When an outbreak such as this occurs, readily available data and knowledge are equally essential to continue the evaluation required to identify the threats and begin outbreak containment activities. From the observation, it is evident that for Bangladesh the number of total confirmed death is increased by a factor of 2.2 in the 5 days (21 deaths on 10 April and 46 deaths on 15 April). In addition, the number of total confirmed case is increased by a factor of 2.1 in the 3 days (482 confirmed case on 12 April and 1012 confirmed case on 15 April). It is also notable that, the number of confirmed cases in the country increased rapidly as the conduction of number of test increased in passage of time. Until 15 April 2020 a total of 14,868-specimen have been tested which was 13.25% higher in comparing with the total test (13,128) performed on 14 April 2020. As of 15 April 2020 69.0% (44/64) district of the country has already been infected with minimum one confirmed COVID-19 case. Until now, Bangladesh has only reported 5.92% (1231/20,804) of confirmed cases in South East Asian Region with a case fatality rate of 5.26 % (50/950). Importantly, regarding the trend of daily confirmed cases in terms of number of days since 100^th^ confirmed cases, Bangladesh (803 on the 7 days of 100 confirmed case) shows the similar trend till now with France (716), USA (754), Pakistan (887) and Thailand (721). However, no countries have the same trend as Bangladesh in terms of days since the fifth confirmed deaths until now. The pandemic has shown us how quickly a new epidemic will catch up and spread. Comprehension is expected with an eruption in clinical and epidemiological information and research. Therefore, the goal of this research is to open up whatever resources we have to help the public health authorities, researchers and clinicians in Bangladesh in mitigating and handling this crisis. The study presented here, using our designed dataset “Bangladesh COVID-19 Daily Dataset” focused on both exploratory and visual exploratory data analysis. All the analysis with the dataset was performed using Tableau Desktop Professional Edition 2020.1.2 64-bit version (https://www.tableau.com/) support on a windows-based local computer platform, using Open Street Map (https://www.openstreetmap.org/) and Map Box (https://www.mapbox.com/) for interactive map visualization in the research laboratory of Dhaka International University (DIU), Bangladesh. Lastly, our research team believes, it is crucial to produce accurate, real-time, and reliable data for emerging disease outbreaks, which will help generate credible information that can reinforce which inform policy taking in public health sector.

## 5. Conclusion

This report analyzed the data focusing on specific criteria including public awareness, case management, infection control, surveillance, quarantine facility and trend analysis with other countries. Consequently, this is, to the best of our knowledge, the first initiative of analyzing and visualizing real-world time series COVID-19 data of Bangladesh in such a way that health professionals and researchers around the globe can grasp its severity clearly. The COVID-19 pandemic has ravaged the lives of people around the world and has become a clinical threat to the general population and healthcare workers around the world. However, awareness on this novel virus (SARS-CoV-2) is still minimal in Bangladesh. This study analyzed the forty (40) days situation since the first COVID-19 confirmed patients reported in the country. Additionally, this exploration also implemented and used the country’s first publicly available daily COVID-19 case dataset. It also illustrated in this study is the day wise case review and assessment of epidemiological evidence with SAARC, SEAR and the top 10 countries worldwide (in terms of deaths). Our research team believes this is an early data analysis and visualization approach of a situation that is changing rapidly across the country. Therefore, as the more we learn about this SARS-CoV-2 virus and its associated outbreaks, the better we can respond. We will continue to monitor and update the epidemiological data of COVID-19 outbreak in the country in upcoming days by through the developed dataset.

## Data Availability

Not Applicable

## Author Contribution

All authors conceptualized and designed the study. SKD and MR had the idea for and designed the study and had full access to all the data in the study and take the responsibility for the data and accuracy of the data analysis with their visualization. URS and AH and contributed to the writing of the article. MR contributed to critical revision of the report. All the visualization and data presentation methods developed by SKD and MR. All authors contributed to data acquisition, data analysis, and reviewed and approved the final version.

## Funding

None

## Declaration of Interests

The authors declare that there are no conflicts of interest.

## Ethical Approval

Not required

## Acknowledgment

The authors would like to acknowledge the Directorate General of Health Services (DGHS) for their regular press release. Also, like to acknowledge Institute of Epidemiology, Disease Control and Research (IEDCR) for their timely situational reports on COVID-19. We are also highly grateful to Ministry of Health and Family Welfare (MoHFW) for making the data publicly available in first place.

## Notes

### Competing Interest Statement

The authors have declared no competing interest.

